# Red blood cell distribution width (RDW) in Hospitalized COVID-19 Patients

**DOI:** 10.1101/2020.06.29.20143081

**Authors:** Preethi Ramachandran, Mahesh Gajendran, Abhilash Perisetti, Karim Osama Elkholy, Abhishek Chakraborti, Giuseppe Lippi, Hemant Goyal

## Abstract

**Introduction:** Coronavirus disease-2019 (COVID-19), caused by severe acute respiratory syndrome coronavirus-2 (SARS-CoV-2), is causing dramatic morbidity and mortality worldwide. The Red Blood Cell Distribution Width (RDW) has been strongly associated with increased morbidity and mortality in multiple diseases.

**Objective:** To assess if elevated RDW is associated with unfavorable outcomes in hospitalized COVID-19.

**Methods:** We retrospectively studied clinical outcomes of hospitalized COVID-19 patients for their RDW values. In-hospital mortality was defined as primary outcome, while septic shock, need for mechanical ventilation, and length of stay (LOS) were secondary outcomes.

**Results:** A total of 294 COVID-19 patients were finally studied. Overall prevalence of increased RDW was 49.7% (146/294). RDW was associated with increased risk of in-hospital mortality (aOR, 4.5; 95%CI, 1.4-14.3) and septic shock (aOR, 4.6; 95%CI, 1.4-15.1) after adjusting for anemia, ferritin, and lactate. The association remained unchanged even after adjusting for other clinical confounders such as age, sex, body mass index, coronary artery disease, hypertension, diabetes mellitus, and chronic obstructive pulmonary disease. No association was found instead with mechanical ventilation and median LOS.

**Conclusion:** Elevated RDW in hospitalized COVID-19 patients is associated with a significantly increased risk of mortality and septic shock.

## 1. Introduction

The ongoing Coronavirus Disease 2019 (COVID-19) pandemic, caused by severe acute respiratory syndrome corona virus-2 (SARS-CoV-2), has started as a cluster of pneumonia-like illness in Wuhan, China, and has now spread all around the world [1]. During the initial outbreak of COVID-19, respiratory involvement was the primary cause of morbidity and mortality. As of June 18, 2020, COVID-19 had infected over 9 million people, causing 450,000 deaths, and numbers continue to rise [2]. As the number of patients increased, other organ system involvement were increasingly recognized [3-8]. The US Center for Disease Control and Prevention (CDC) emphasizes that patients with preexisting conditions such as advanced age (65 years or older) or pathologies like cardiovascular disease, diabetes, cancer, hypertension, chronic obstructive pulmonary disease (COPD) are at higher risk of COVID-19-associated morbidity and mortality [9].

It is now unquestionable that early identification and prompt management of COVID-19 could prevent or delay the onset of life-threatening complications. In addition to preexisting conditions, the use of prognostic laboratory biomarkers may be of great clinical significance for identifying patients at higher risk of worse progression. Similar to other viral illnesses, COVID-19 is associated with leukopenia, lymphopenia, and elevated values of many traditional inflammatory biomarkers [10, 11].

RDW is an inexpensive measure of erythrocyte size variation, and it can be used in the differential diagnosis of hematological disturbances such as iron deficiency anemia and bone marrow dysfunction [12]. This parameter is calculated as the standard deviation (SD) of red blood cell (RBC) volume, divided by the mean corpuscular volume (MCV), thus providing a quantitative estimation of anisocytosis. Alteration of erythropoiesis can result in extensive heterogeneity of RBC size, which can provide indirect evidence of existing and ongoing pathological changes. In inflammatory states, the turnover of the RBC is decreased, with a simultaneous increase of inflammatory cell turnover (e.g., leukocytes and platelets) in the attempt to counteract the infection [13]. Due to these changes, RDW is usually elevated in conditions such as advanced age, diabetes, cardiovascular diseases, gastrointestinal disorders, infections, and a vast array of infections and inflammatory states [14-18].

As the pathogenesis of COVID-19 involves both infection and inflammation, the RDW measured at hospital admission could potentially be considered a reliable index for identifying patients at higher risk of an unfavorable outcome [19]. Therefore, this study was aimed at exploring whether RDW value upon admission may predict clinical progression in patients hospitalized for COVID-19.

## 2. Methods

This retrospective descriptive study was carried out from a single tertiary care academic medical center in New York City. The study was approved by the institutional review board.

The initial study population consisted of all consecutive adult patients admitted to the hospital with confirmed SARS-CoV-2 infection from January 20, 2020, to April 25, 2020. The study cohort was stratified into two groups according to the RDW value (elevated, cases; normal, controls). The RDW is measured using the Beckman Coulter analyzer. Elevated RDW was defined as a value >14.6%, which is the upper limit of the healthy adult reference interval in the local institution. Inclusion criteria included all adult patients (age ≥ 18 years) hospitalized for COVID-19, thus testing positive for SARS-CoV-2 on the nasopharyngeal swab, according to current guidelines [20]. Exclusion criteria were-individuals who were not hospitalized or treated on an ambulatory basis, age < 18 years, pregnancy, non-availability of nasopharyngeal testing, lack of clinical or laboratory information on disposition and/or mortality data anemia was defined as a hemoglobin value <130 g/L in males and <120 g/L in females, respectively.

Information on patient demographics, presenting symptoms, comorbidities, home medications, and initial laboratory tests were obtained at hospital admission. In-hospital mortality from any cause was defined as the primary outcome in this study, whilst septic shock, need for mechanical ventilation, and in-hospital length of stay (LOS) were secondary outcomes. Statistical analysis was performed using IBM SPSS software version 26 (SPSS Inc, Armonk, NY). Descriptive summary statistics are shown as median with interquartile range (IQR) for continuous variables since most were not normally distributed, and frequencies with percentages for categorical variables. Categorical and continuous variables were tested for statistical significance using chi-square tests and t-tests, respectively. We performed two models of multivariable logistic regression analyses. In the first model, we included variables with P <0.05 from univariate analysis. In the second model, we also included other clinically relevant variables such as age, sex, body mass index (BMI), coronary artery disease (CAD), hypertension, diabetes mellitus, and chronic obstructive pulmonary disease (COPD). The results of the regression models were provided as adjusted odds ratio (aOR) and its 95% confidence interval (CI). The receiver operating characteristic (ROC) curve was used for exploring the discriminant power of the model.

## 3. Results

The initial study population consisted of 300 patients hospitalized for COVID-19, six of whom ought to be excluded due to undefined results of SARS CoV-2 testing and lack of key outcome variables. Therefore, a final number of 294 patients composed the final study population (Figure 1). The prevalence of elevated RDW values was found to be 49.7% (146/294).

**Figure 1:**
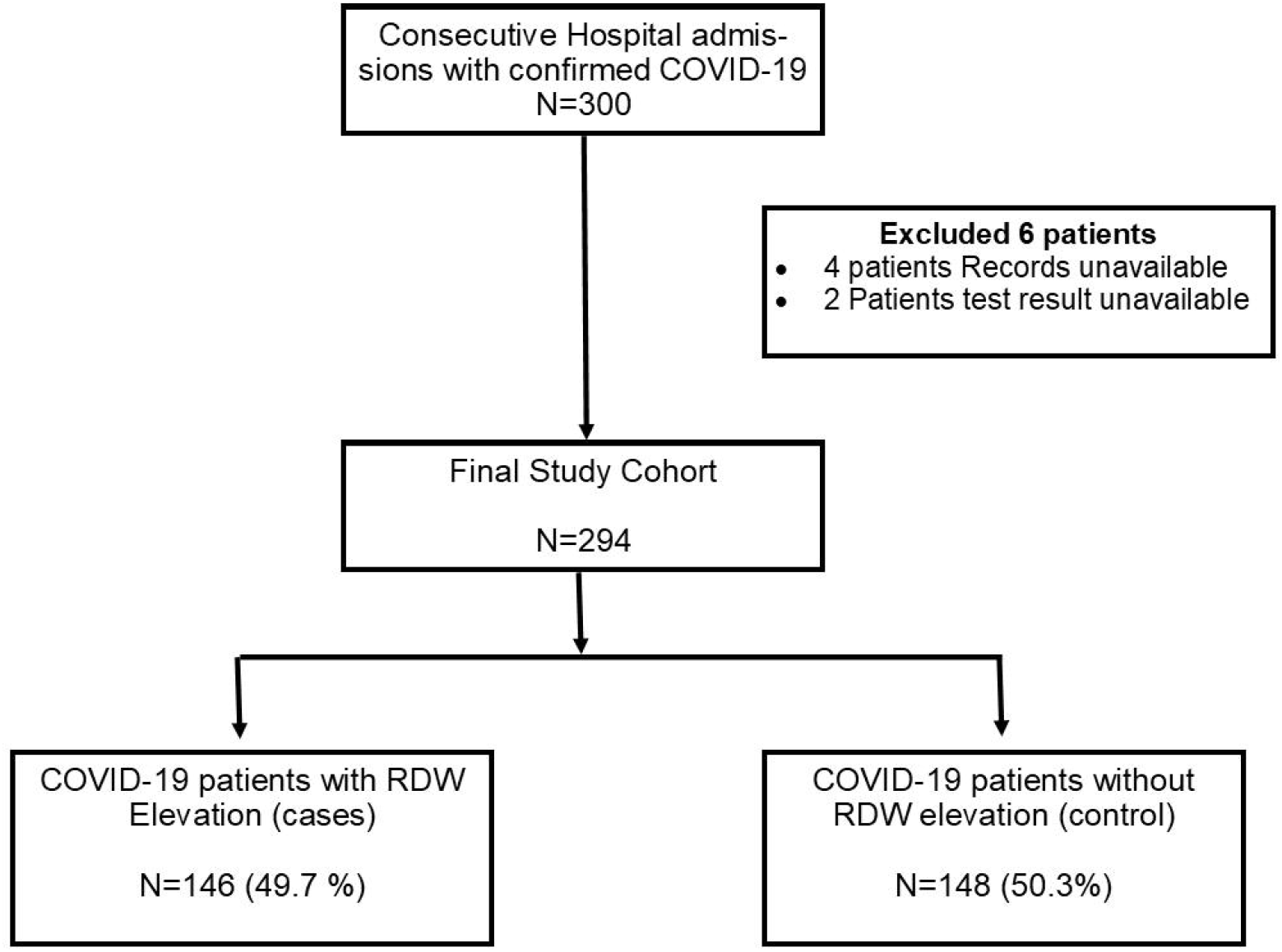
Study Flowchart Footnotes: COVID-19: Coronavirus Disease 2019; RDW: Red blood cell distribution width

### 3.1 Baseline demographics

Baseline demographic data and comorbidities in patients with or without elevated RDW value are summarized in Table 1. Patients with elevated RDW were older (median age, 66 vs. 63 years; p=0.02), with female predominance (52% vs. 38%; p=0.02). Patients with elevated RDW value had also a higher prevalence of hypertension (79.5% vs. 62.8%; p=0.002), CAD (19.9% vs. 10.8%; p=0.04) and COPD (13% vs. 5.4%; p=0.03).

**Table 1:**
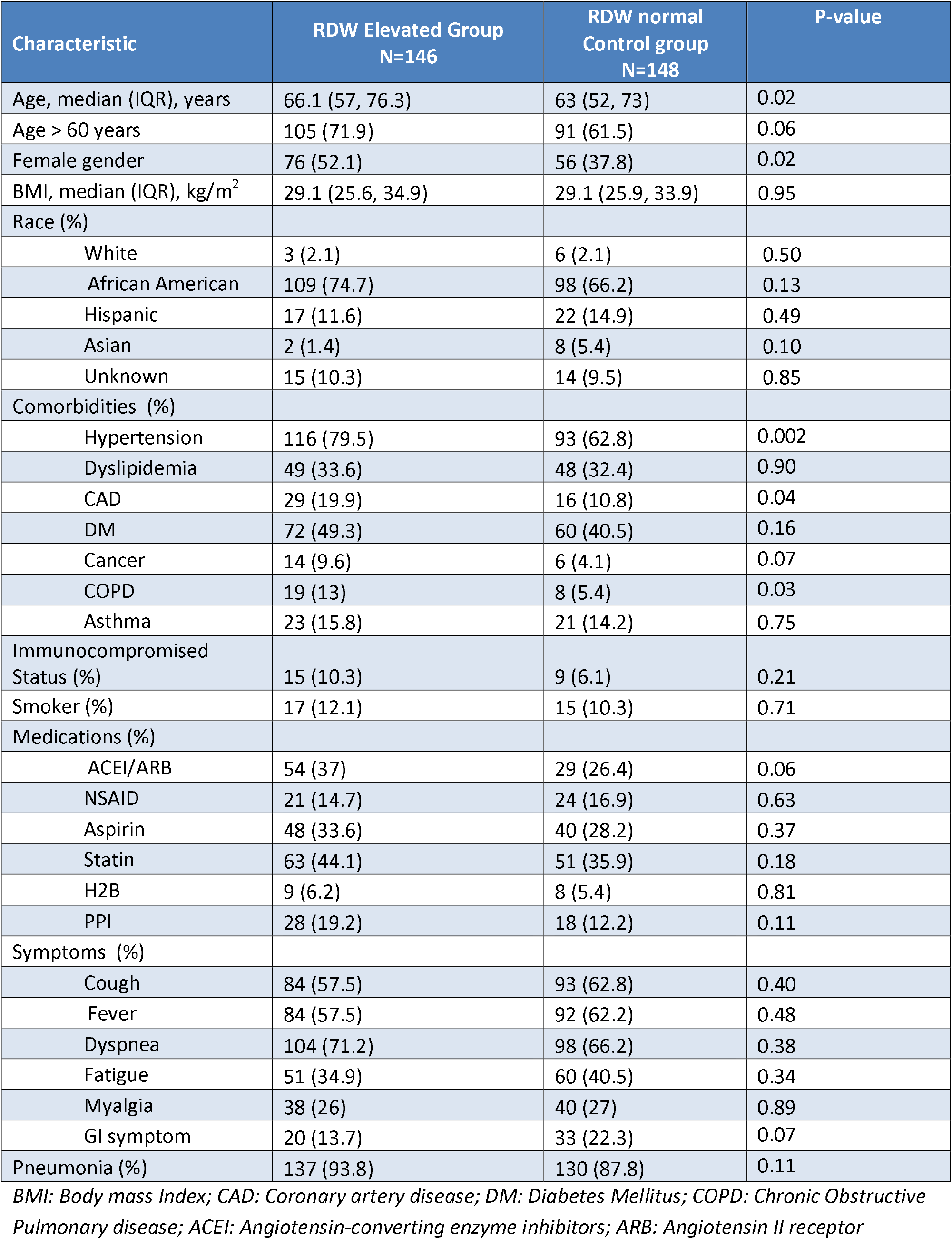
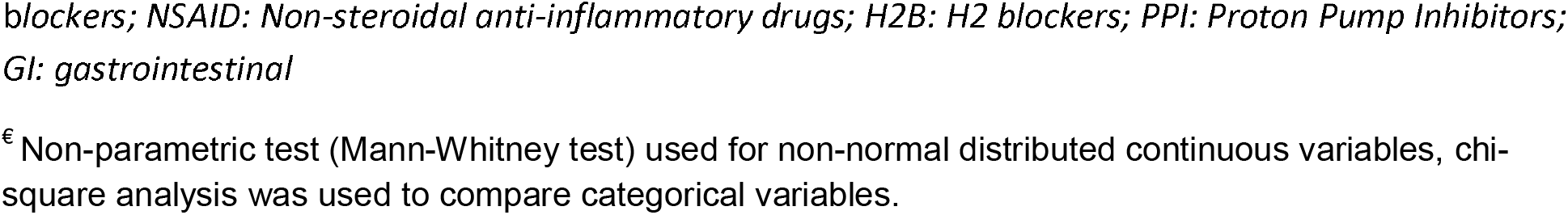
Baseline demographics of the study population ^€^.

| Characteristic | RDW Elevated Group<br>N=146 | RDW normal<br>Control group<br>N=148 | P-value |
| --- | --- | --- | --- |
| Age, median (IQR), years | 66.1 (57, 76.3) | 63 (52, 73) | 0.02 |
| Age > 60 years | 105 (71.9) | 91 (61.5) | 0.06 |
| Female gender | 76 (52.1) | 56 (37.8) | 0.02 |
| BMI, median (IQR), kg/m<sup>2</sup> | 29.1 (25.6, 34.9) | 29.1 (25.9, 33.9) | 0.95 |
| Race (%) |  |  |  |
| White | 3 (2.1) | 6 (2.1) | 0.50 |
| African American | 109 (74.7) | 98 (66.2) | 0.13 |
| Hispanic | 17 (11.6) | 22 (14.9) | 0.49 |
| Asian | 2 (1.4) | 8 (5.4) | 0.10 |
| Unknown | 15 (10.3) | 14 (9.5) | 0.85 |
| Comorbidities (%) |  |  |  |
| Hypertension | 116 (79.5) | 93 (62.8) | 0.002 |
| Dyslipidemia | 49 (33.6) | 48 (32.4) | 0.90 |
| CAD | 29 (19.9) | 16 (10.8) | 0.04 |
| DM | 72 (49.3) | 60 (40.5) | 0.16 |
| Cancer | 14 (9.6) | 6 (4.1) | 0.07 |
| COPD | 19 (13) | 8 (5.4) | 0.03 |
| Asthma | 23 (15.8) | 21 (14.2) | 0.75 |
| Immunocompromised<br>Status (%) | 15 (10.3) | 9 (6.1) | 0.21 |
| Smoker (%) | 17 (12.1) | 15 (10.3) | 0.71 |
| Medications (%) |  |  |  |
| ACEI/ARB | 54 (37) | 29 (26.4) | 0.06 |
| NSAID | 21 (14.7) | 24 (16.9) | 0.63 |
| Aspirin | 48 (33.6) | 40 (28.2) | 0.37 |
| Statin | 63 (44.1) | 51 (35.9) | 0.18 |
| H2B | 9 (6.2) | 8 (5.4) | 0.81 |
| PPI | 28 (19.2) | 18 (12.2) | 0.11 |
| Symptoms (%) |  |  |  |
| Cough | 84 (57.5) | 93 (62.8) | 0.40 |
| Fever | 84 (57.5) | 92 (62.2) | 0.48 |
| Dyspnea | 104 (71.2) | 98 (66.2) | 0.38 |
| Fatigue | 51 (34.9) | 60 (40.5) | 0.34 |
| Myalgia | 38 (26) | 40 (27) | 0.89 |
| GI symptom | 20 (13.7) | 33 (22.3) | 0.07 |
| Pneumonia (%) | 137 (93.8) | 130 (87.8) | 0.11 |

BMI: Body mass Index; CAD: Coronary artery disease; DM: Diabetes Mellitus; COPD: Chronic Obstructive Pulmonary disease; ACEI: Angiotensin-converting enzyme inhibitors; ARB: Angiotensin II receptor
blockers; NSAID: Non-steroidal anti-inflammatory drugs; H2B: H2 blockers; PPI: Proton Pump Inhibitors; GI: gastrointestinal
€ Non-parametric test (Mann-Whitney test) used for non-normal distributed continuous variables, chi-square analysis was used to compare categorical variables.

### 3.2 Laboratory data

Laboratory data at the time of admission in patients with or without elevated RDW value are summarized in Table 2. The prevalence of anemia was higher in patients with increased RDW value than in those without (58.2% vs. 33.1%; p<0.001). Elevated lactate values were also more frequent in patients with increased RDW value than in those without (48.3% vs. 33.1%; p= 0.018). The rate of abnormal values of lymphocyte count, platelet count, D-dimer, C reactive protein (CRP), aminotransferases, bilirubin, and alkaline phosphatase did not differ between the two cohorts of patients.

**Table 2:** **Laboratory data in hospitalized COVID-19 patients according to their red blood cell distribution width (RDW) value**

| Characteristic | RDW Elevated Group<br>N=146 | RDW normal group<br>N=148 | P-value |
| --- | --- | --- | --- |
| Anemia | 85 (58.2) | 49 (33.1) | <0.001 |
| Elevated Ferritin | 33/44 (75) | 43/47 (91.5) | 0.048 |
| Elevated D-dimer | 28/34 (82.4) | 30/35 (85.7) | 0.75 |
| Leukocytosis | 39 (26.7) | 32 (21.6) | 0.34 |
| Lymphopenia | 88 (60.3) | 81 (54.7) | 0.35 |
| Thrombocytopenia | 24 (16.4) | 37 (25) | 0.08 |
| Elevated Creatinine | 62 (42.5) | 48 (32.9) | 0.17 |
| Hypoalbuminemia | 43 (29.5) | 37 (25.2) | 0.43 |
| Elevated Creatinine Phosphokinase | 9/69 (13) | 11/86 (12.8) | 1.00 |
| Elevated Lactate | 57/118 (48.3) | 40/121 (33.1) | 0.018 |
| Elevated LDH | 124/124 (100) | 128/129 (99.2) | 1 |
| Elevated CRP | 67 /117 (57.3) | 59/121 (48.8) | 0.19 |
| Elevated AST | 65 (44.8) | 74 (50.3) | 0.35 |
| Elevated ALT | 21 (14.6) | 30 (20.4) | 0.22 |
| Elevated Bilirubin | 19 (13) | 15 (10.2) | 0.47 |
| Elevated Alkaline Phosphatase | 25 (17.2) | 16 (10.9) | 0.13 |

LDH: Lactate Dehydrogenase; CRP: C-reactive protein; AST: Aspartate aminotransferase; ALT: Alanine aminotransferase

### 3.3 Outcomes

The outcomes of the study are shown in Table 3. The COVID-19 patients with elevated RDW value had a higher frequency of in-hospital mortality compared to those with normal RDW value, but the difference was not statistically significant (23.3% vs. 15.1%, P=0.1). However, after adjusting for lactate, ferritin, and anemia, elevated RDW was found to be significantly associated with a higher risk of in-hospital mortality (aOR 4.5, 95% CI 1.44-14.28; p=0.01). The incidence of shock was also higher in the elevated RDW group (37.7% vs. 25.7%; p=0.03) compared to patients with normal RDW values. Elevated RDW was associated with a 4.6-fold higher odds of shock (aOR 4.6, 95% CI 1.43-15.08; p=0.01) after adjusting for confounders. No association with RDW value was found for the need for mechanical ventilation or in-hospital (LOS).

**Table 3:** **Outcomes data on hospitalized COVID-19 patients according to their red blood cell distribution width (RDW) value**

| Characteristic | RDW Elevated Group<br>N=146 | RDW normal group<br>N=148 | P-value |
| --- | --- | --- | --- |
| In-hospital Mortality | 34 (23.3) | 22 (15.1) | 0.10 |
| Shock | 55 (37.7) | 38 (25.7) | 0.03 |
| Mechanical Ventilation | 29 (19.9) | 20 (13.5) | 0.19 |

**Table 4:** **Multivariable Logistic Regression Analysis of elevated red blood cell distribution width (RDW) value**

| Variables | Adjusted Odds Ratio (aOR) with 95%<br>Confidence Interval (CI) | P-value |
| --- | --- | --- |
| **Model 1<sup>€</sup>** |  |  |
| Death | 4.5 (1.4 – 14.3) | 0.010 |
| Septic Shock | 4.6 (1.4 – 15.1) | 0.010 |
| Mechanical Ventilation | 2.1 (0.7 – 6.5) | 0.191 |
| **Model 2<sup>£</sup>** |  |  |
| Death | 5.5 (1.3 – 22.8) | 0.018 |
| Septic Shock | 5.4 (1.3 – 21.1) | 0.016 |
| Mechanical Ventilation | 1.4 (0.4 – 4.9) | 0.576 |

<sup>€</sup>Model 1: Adjusted for Anemia, Elevated Ferritin and elevated Lactate
<sup>£</sup>Model 2: Adjusted for Age, Gender, Body mass index, coronary artery disease, Hypertension, Diabetes Mellitus, Chronic Obstructive Pulmonary Disease, Anemia, Elevated Ferritin and elevated Lactate

### 3.4 Analysis by ROC

The ROC area under the curve (AUC) was found to be 0.85 and 0.77 for predicting mortality (Figure 2A) and septic shock (Figure 2B), respectively. It had a sensitivity of 70% in predicting both mortality and septic shock.

**Figure 2 (A):**
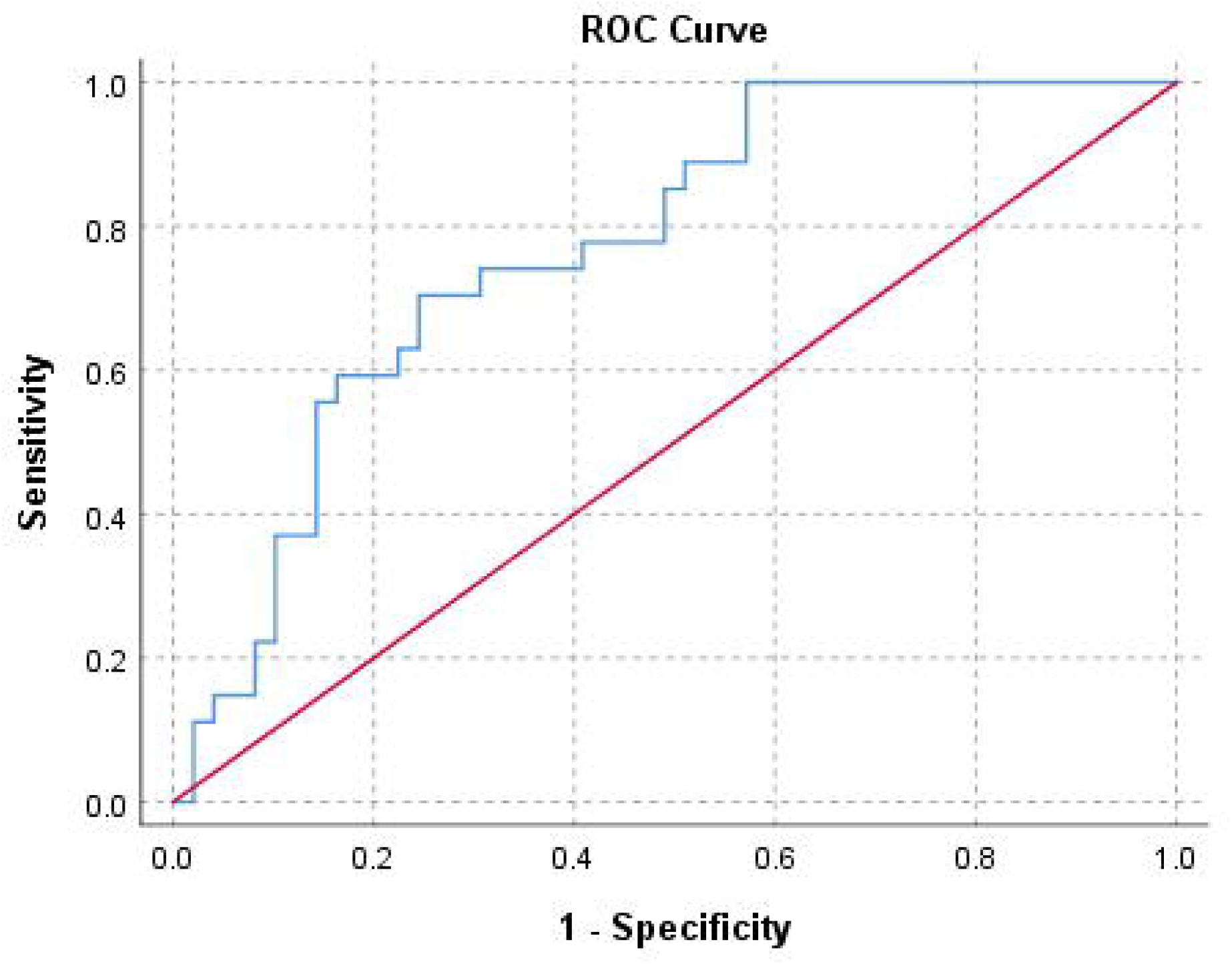
Receiver Operator Curve (ROC) of red blood cell distribution width (RDW) values in coronavirus disease 2019 (COVID-19) patients for predicting mortality

**Figure 2 (B):**
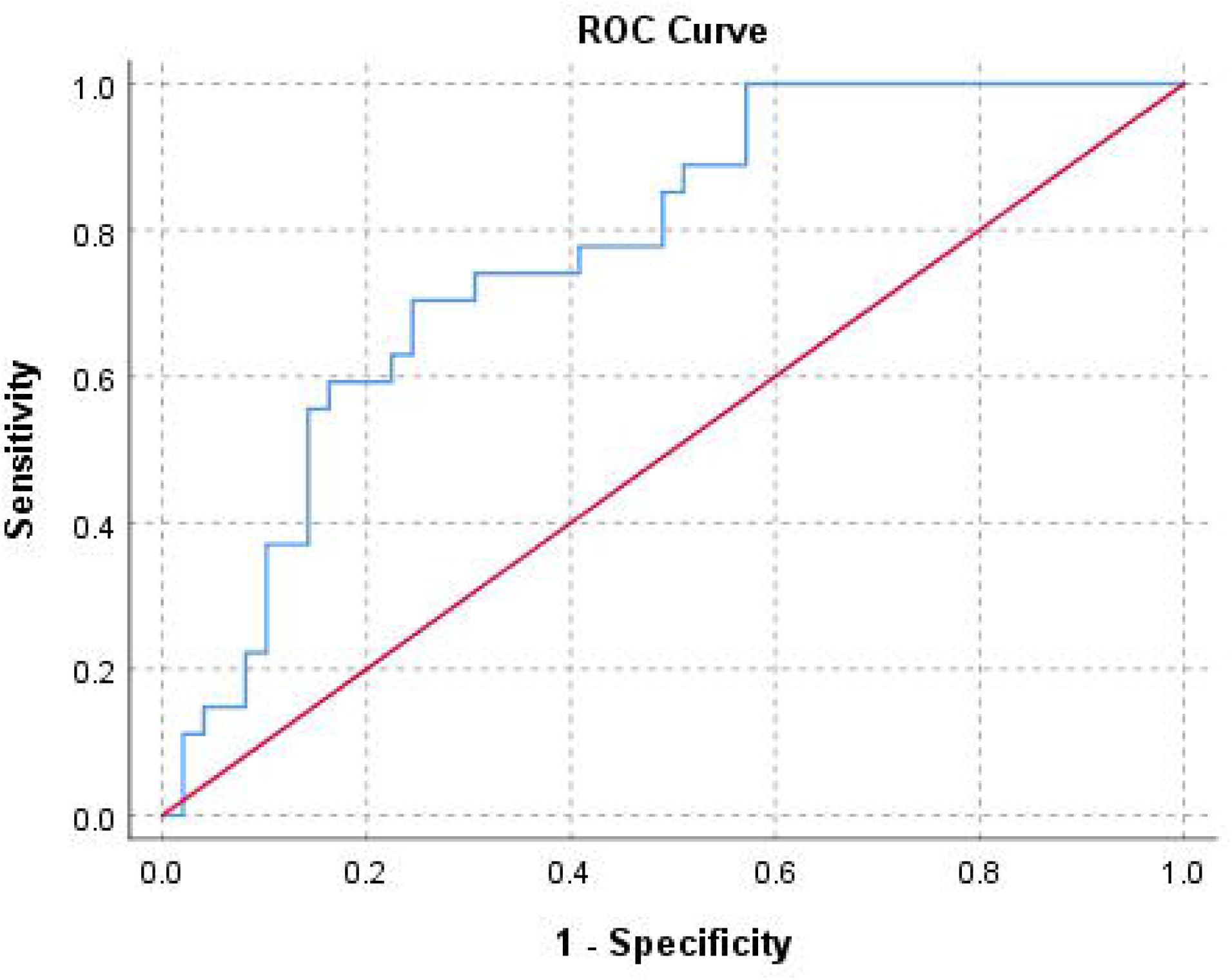
Receiver Operator Curve (ROC) of red blood cell distribution width (RDW) values in coronavirus disease 2019 (COVID-19) patients for predicting septic shock

## 4. Discussion

Based on our study, nearly half (49.7%) of patients hospitalized for COVID-19 were found to have elevated RDW values at presentation. Patients with increased RDW, and thereby more significant anisocytosis, had increased risk of in-hospital mortality and septic shock compared to those with normal values. These findings remained significant after adjusting for potential confounders. On the other hand, we failed to find a significant association between RDW and the need for mechanical ventilation and LOS.

RDW increases with age, especially among females [21]. This is probably due to decreased RBC deformability developing in parallel with aging. Similarly, RDW is identified as an important risk factor in the diagnosis and prognosis of patients with cardiovascular disease (CVD) [12]. While the precise mechanism remains speculative, a combination of RDW and validated cardiac markers can help to identify patients with CVD earlier, thus allowing them to establish more appropriate and targeted management. Our study showed a similar prevalence of higher RDW with older age and female sex among cases compared to controls. Furthermore, the presence of hypertension and CAD was higher in patients with increased RDW [22, 23], and this is in keeping with earlier studies and is also representative of populations with preexisting health conditions [13, 14, 21].

The precise mechanism by which RDW elevation develops in COVID-19 patients is unclear. Prior studies showed an association of elevated RDW with increased inflammatory markers, oxidative stress with impaired iron metabolism, which would ultimately promote RBC apoptosis, and variance in their morphology [13, 24, 25]. COVID-19 patients are known to have a significant inflammatory response, which can lead to multiorgan failure. It is unclear if increased RDW in COVID-19 patients is due to this inflammatory response [26]. It is well known that SARS-CoV-2 enters human cells via angiotensin-converting enzyme 2 (ACE-2) [27]. The expression of ACE-2 varies significantly among organs and tissues, and this would contribute to explaining the different degrees of the vulnerability of host cells to viral entry and cytopathic effects [28]. The use of medications such as ACEI/ARB in our study was higher in cases compared to controls (37% vs. 26.4%; p=0.06) but did not reach statistical significance. It can, at least in part, explain the gastrointestinal manifestations such as nausea, vomiting, diarrhea and abnormal liver tests in patients with enhanced RDW values.

Higher admission RDW may reflect the presence of ongoing chronic and severe inflammation. Some patients with COVID-19 develop a cytokine storm syndrome, characterized by overproduction of early response pro-inflammatory cytokines such as tumor necrosis factor, interleukin [IL]-6, and IL-1β [26]. It can lead to increased vascular permeability, hyper-inflammation, and loss of procoagulant-anticoagulant balance [26]. Our study showed an increased level of lactate in patients with RDW values above the normal threshold, thus correlating with the increased incidence of shock and mortality. The increased RDW was found to be independently associated with a higher incidence of shock and mortality after adjusting for all the confounding variables. Whether higher RDW can predict rates of altered hemodynamics and early features of shock in these patients is unclear and remains to be studied.

RDW has been previously studied in patients with severe sepsis and septic shock. Wang et al. noted that higher levels of RDW was an independent predictor of in-hospital mortality in elderly patients with sepsis, and could potentially be used as a reliable biomarker for predicting clinical outcomes [29]. RDW has also been used as a prognostic biomarker for predicting 28-day mortality in patients with severe sepsis and septic shock [30, 31]. Recently Foy et al. showed that elevated RDW at diagnosis, as well as its increase during the hospitalization, was associated with an increased risk of mortality in patients with COVID-19 [32], thus supporting our evidence. Nonetheless, the need for mechanical ventilation and the LOS did not differ between cases and controls, and this probably attributable due to adequate resuscitative efforts enabled by an early risk stratification. In addition, other markers of inflammation such as D-dimer, leukocytosis, C-reactive protein (CRP) did not differ significantly according to the RDW threshold, thus paving the way to further studies aimed at precisely establishing the role of anisocytosis in the pathogenesis of COVID-19.

## 5. Conclusion

Elevated RDW is a common finding in patients hospitalized for COVID-19. The preliminary findings of this study show that elevated RDW at admission is present in almost half of patients, and independently predicts shock and mortality but not LOS or need for mechanical ventilation.

## Data Availability

Anonymous data can be arranged if needed

## Acknowledgments

*None*

